# Prevalence of Parkinson’s disease in Lagos, Southwestern Nigeria: a descriptive community-based study from the Transforming Parkinson’s Care in Africa (TraPCAf) project

**DOI:** 10.64898/2026.06.27.26356731

**Authors:** NU Okubadejo, OO Ojo, A Ogunyemi, OP Agabi, A Oyeleye, F Nwaokorie, R Anyanwu, D Ezuduemoih, O Ibode, S Chabiri, O Madueke, E Ikwenu, R Morton, SJ Urasa, MCJ Dekker, CL Dotchin, N Fothergill-Misbah, M Cham, A Akpalu, RW Walker, the TraPCAf consortium

**Author notes:** **CORRESPONDENCE** Njideka U. Okubadejo Neurology Unit, Department of Medicine, Faculty of Clinical Sciences, College of Medicine, University of Lagos, Lagos State, Nigeria. **DISCLOSURES**. NUO – honoraria and travel support (International Parkinson’s and Movement Disorders Society [MDS]); grants (Michael J Fox Foundation [MJFF], United Kingdom National Institute for Health and Care Research [UK NIHR). OOO – honoraria and travel support (MDS, MJFF), Grants (UK NIHR).

## Abstract

**Background:** The global burden of Parkinson’s disease (PD) has increased substantially over recent decades, driven by population ageing and rising age-standardized prevalence. In Africa, accurate estimates remain limited due to a lack of recent, methodologically robust population-based studies.

**Objectives:** To determine the current age-standardized and sex-specific prevalence rates of PD in Nigeria.

**Methods:** We conducted a 2-stage, cross-sectional population-based door-to-door survey among adults aged ≥18 years in two densely populated urban local government areas in Lagos State, Nigeria, between April 1, 2024 and January 31, 2025. The first stage involved a household census and screening for parkinsonism using a standardized screening tool. The second stage consisted of in-person clinical assessment and diagnostic confirmation by physicians using established clinical diagnostic criteria. Crude and age-standardized prevalence rates (to the World Health Organization World Standard and European Standard Populations) were calculated.

**Results:** 31,009 individuals (52.7% female) from 13,222 households were surveyed, and 70 persons were diagnosed with PD. The crude prevalence ratio was 225.7 per 100,000, with higher prevalence in males (53/14658, 361.6) than females (17/16,351, 104.0). The age-standardized prevalence rate (95% confidence interval) was 193 per 100,000 (150 –245) (females: 86 (50 – 137); males: 277 (207 – 362)), and increased with advancing age. The diagnostic gap (previously undiagnosed) was 60.0% (42/70). Treatment gap (never treated) was 44/70 (62.9%).

**Conclusions:** The age-standardized prevalence of PD is higher than previously reported in sub-Saharan Africa. These findings provide contemporary data to inform updated estimates of disease burden and support health systems planning.

## INTRODUCTION

Parkinson’s disease (PD) is the second most common neurodegenerative disorder worldwide. The Global Burden of Disease (GBD) Study first highlighted PD as the fastest growing neurological disorder between 1990 and 2016. (1) This increase in burden was documented across most world regions, including sub–Saharan Africa (SSA), and was suggested to be attributable to a combination of factors including an increase in numbers of the highest at risk population (older people), longer disease duration, and environmental factors. (2) This position is reiterated in the GBD 2021 systematic analysis of disorders affecting the nervous system (1990-2021). (3) In 2021, the percentage change in prevalence counts for PD (from 1990) was 273.9%, exceeding that for Alzheimer’s disease and other dementias (ADOD) (160.8%) and stroke (86.1%). Similarly, the percentage change in age-standardized prevalence rate per 100,000 people for PD (1990-2021) was 60.7% in contrast to ADOD (3.2%) and stroke (minus 8.5%), and all neurological conditions combined (1.5%). (3)

Whereas substantial epidemiological data exists from high-income countries, there is a paucity of community-based studies from low-and middle-income countries (LMICs), particularly in Africa. This gap impacts negatively on the availability of evidence to substantiate the burden of PD in a continent that has undergone demographic transitions (including population ageing) which may shape the disease burden differently. There are limited epidemiologic data on PD from LMICs on account of challenges conducting community-based surveys, difficulty accessing rural areas, and limited hospital registers. (4) The handful of published epidemiological studies have demonstrated varying methodological quality, were published more than a decade ago, and may not represent the current scenario. Africa is experiencing a demographic shift with an increase in the number of older persons aged 60 and above, a projected tripling of the numbers by 2050, outstripping growth rates of any other world region. (5)

The World Health Organization (WHO) technical brief addressing the public health approach to PD specifically identifies and highlights the crucial role of epidemiological PD research as one of the key actions to address the burgeoning burden of PD. (6) This data serves as crucial evidence for advocacy, formulating policies and implementing health systems restructuring to bridge the gaps in access to care for people with PD. The GBD projections and estimates of PD burden in Africa have been constrained by the lack of methodologically robust and representative studies to accurately quantify prevalence and disability-adjusted life years.

The Transforming Parkinson’s Care in Africa (TraPCAf) project is a multi-methodology project that includes prevalence studies across four locations in Africa, aimed at defining the current burden of PD by providing updated prevalence data. (7) As part of this initiative, we conducted a descriptive, community-based study to estimate the prevalence of PD in Lagos. By generating robust local data, this study aims to contribute to the global epidemiological understanding of PD, inform health policy, and support the development of context-appropriate strategies for diagnosis, management, and care delivery in Nigeria and beyond.

## METHODOLOGY

### Study design

This descriptive epidemiological (prevalence) study was conducted in two local government areas (LGAs) in urban Lagos, Southwest Nigeria. The study is one of four community-based door-to-door studies conducted in four African countries (Nigeria, Kenya, Ghana and Tanzania) participating in the TraPCAf project. (7) This study was conducted and reported in accordance with the Standards of Reporting of Neurological Disorders (STROND) guideline. (8, 9) We utilized a 2-stage cross-sectional population-based door-to-door study, conducted between April 1, 2024, and January 31, 2025. In Stage 1 (April 1, 2024 – June 6, 2024), we undertook a household census of persons aged 18 years and above in the communities and screened for features of parkinsonism using a 5-item PD screening tool. In Stage 2 (April 8, 2024 – January 31, 2025) in-person verification of the screening survey results (proportion of randomly selected screen negative, and all screen positives) and verification of probable parkinsonism and PD was conducted.

### Ethical approval

Approval of the study protocol was obtained from the Health Research Ethics Committee (HREC) of the Lagos University Teaching Hospital (Approval number ADM/DSCST/HREC/APP/5520). Written informed consent was obtained from each study participant.

### Setting

The study was conducted in 2 contiguous, demographically delineated urban LGAs (Surulere and Mushin) in Lagos State, Nigeria. Lagos State is in the South-West zone of Nigeria and is administratively divided into 20 constitutionally recognized LGAs (16 urban and 4 peri-urban). According to the National Population Commission (NPC), Lagos State had a projected population of 13,491,804 in 2022. (10) Surulere and Mushin LGAs had projected populations (area) of 744,400 (20.01km^2^) and 935,400 (17.01 km^2^) respectively in the same time reference (2022). (11) LGAs are designated into Census Enumeration Areas (EAs) by the NPC. Each EA is comprised of the equivalent of 1,000 households and presumed to hold 4 occupants each – 2 adults and an average of 2 children. The EAs are geographically mapped, with clear physical coordinates described by street names and house or plot numbers. Maps of each EA were obtained from the NPC. In addition, re-verification of street locations and boundaries was enabled via publicly available satellite maps. Both LGAs are mixed-income, predominantly residential, accessible by land, and have been the site of previous epidemiological surveys. Granular details of internal migration (in and out) are not available for LGAs. However, due to Lagos’s commercial location, internal migration inflow is considered as very high (mainly youth seeking employment/livelihood), while out-migration is moderate (to suburban areas and other states, driven by relatively higher living costs within metropolitan Lagos). Study participants had to have been resident in the designated location as at January 1, 2024 (3 months prior to the study).

Community entry permissions were obtained from the Lagos State Primary Health Care Board, LGA authorities, and the community gatekeepers (including the Community Development Associations). The Medical Officer of Health and Health educators at each LGA facilitated the recruitment of field workers resident within the LGA, specifying a minimum educational level of post-secondary (i.e., ≥14 years of education). The recruited field workers were trained (in March 2024) on recognition of symptoms of parkinsonism. The training included videos and real live patients, administration of the electronic questionnaire designed for the household census/survey, and the individual-level screening questionnaire (described below). The assignment of EAs was strategic to ensure that sampling covered the breadth of each LGA.

### Sample size and sampling method

The minimum sample size for the study was computed using a generic sample size calculator with the following presumptions: prevalence of 0.15% (global pooled prevalence of 1.51 per 1,000) (12), 95% confidence, precision of 30% of the prevalence i.e., ±0.00045 (recommended for rare conditions). This yielded a minimum sample size of 28,400, to which a 5% attrition rate was added, bringing the total to 29,820.

Data was collected using a cluster sampling technique. Clusters were defined using census EAs within the 2 LGAs, followed by a random selection of primarily residential streets. In each cluster, a door-to-door survey was utilized with paired field workers assigned a specified number of households to survey per week.

### Source population and study instruments (Stage 1)

For Stage 1 (household survey and individual screening), we designed the electronic instrument comprised of 2 questionnaires in KoboToolbox ®. In summary, the instrument included an interviewer-administered Household Survey that captured the field workers identification, consent to participate, unique household ID (and a linked hard-copy household address log), household contact person, mobile telephone numbers, number of adults (aged ≥ 18 years) resident in the household (including their study-assigned identifiers, date of birth, and sex).

The second questionnaire was an individual-level questionnaire that captured information from each adult resident in the home (at the same time as the census, or, when not physically present, at a second scheduled visit). The individual forms documented identifiers, sex, date of birth, duration in the residence, the responses to the screening questions, and follow-up questions depending on the screening responses. The 5-item screening questionnaire (Figure 1) was adapted (via a series of consensus meetings of the participating investigators) from that used by Dotchin *et al* which was adapted from previously developed screening questionnaires. (13) The modifications were intended to ensure the questions were contextually and culturally relevant and sensitive, while retaining the core symptoms of parkinsonism typically recognizable symptomatically (slowness, limb tremor, and gait difficulties).

**Figure 1.**
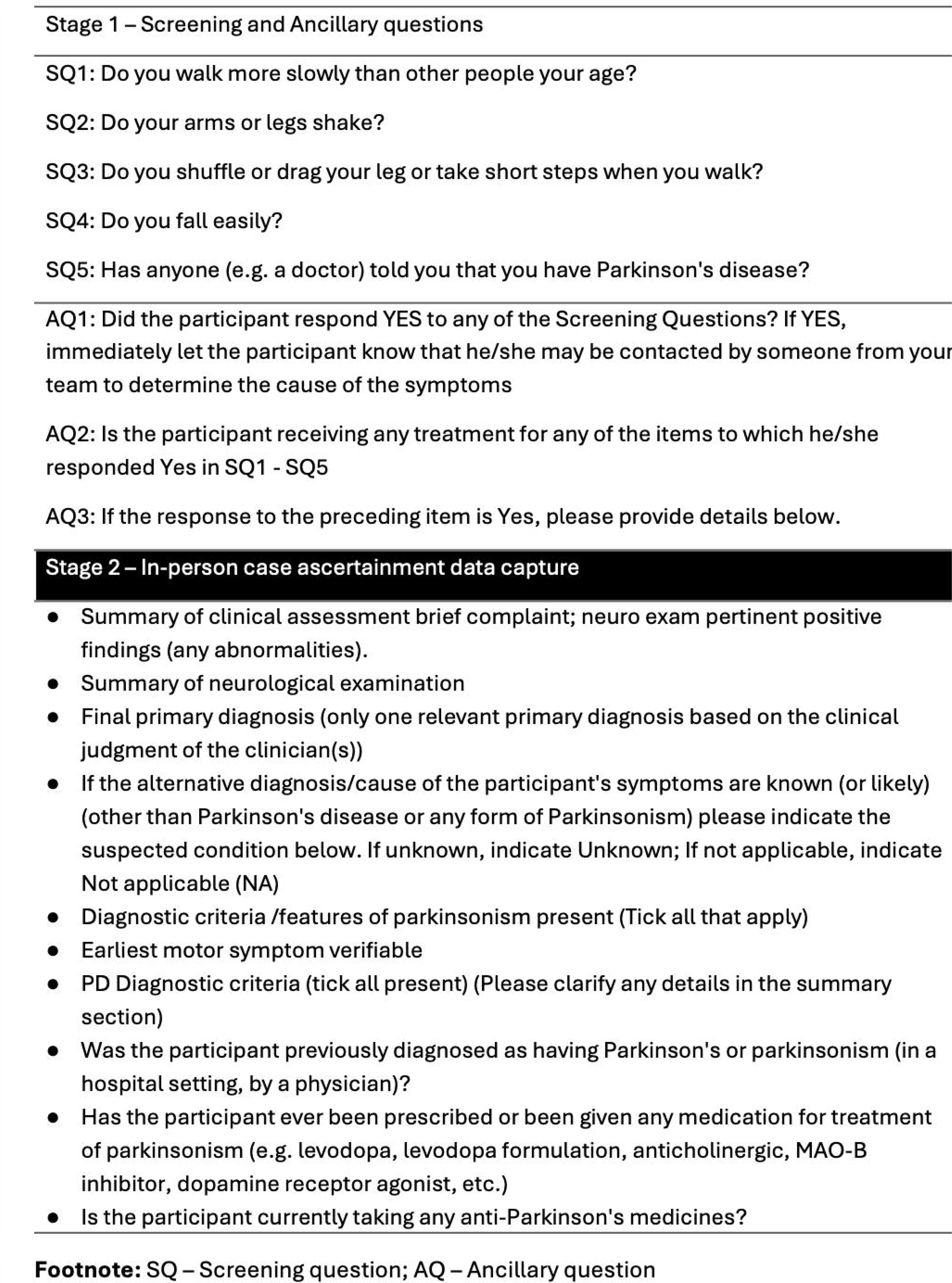
Stage 1 and Stage 2 survey questions and data capture

### Case ascertainment (Stage 2)

Stage 2 was commenced within 2 weeks of Stage 1 to limit loss to follow-up/attrition of the participants and was designed to verify the cause of the positive screen, the presence of parkinsonism, ascertain the diagnosis of PD, and determine the sensitivity and specificity of the screening instrument used in Stage 1. Stage 2 was conducted by a team of 7 study neurologists and 3 medical officers (physicians) trained to recognize parkinsonism and PD. Data generated from the individual screening was reviewed weekly to identify all who screened positive, and in-person evaluation was then scheduled. The in-person assessments included a neurological evaluation and structured documentation of the findings for all participants who screened positive.

Data was captured electronically via a questionnaire designed in KoboToolbox ®.

In summary, the data capture included date/time stamp, identity of physician or neurologist, unique household ID, re-confirmation of consent, identifiers, date of birth and sex, contact information, and details of neurological evaluation, including diagnostic criteria and final primary diagnosis (Figure 1). The diagnosis of PD was based on fulfilment of the United Kingdom Parkinson’s Disease Brain Bank (UKPDBB) criteria (except non-exclusion due to a positive family history; including possible and probable PD by Gelb’s criteria). (14, 15) Additional clinical data related to PD were collected as part of participant inclusion into the TraPCAf work package exploring the clinical profile of PD in Africans. ( 7 )

### Sensitivity and specificity of the screening questionnaire

The utility of the screening questionnaire was evaluated in this study. We selected all the screen positive participants (affirmation of 1 of the main survey questions screening for parkinsonism, n=569) and 599 randomly selected participants who screened negative. These participants were recontacted and underwent an in-person re-evaluation by a study physician using the instrument described for case ascertainment in Stage 2 (and as part of the Stage 2 evaluation). The sensitivity and specificity were calculated using the numbers of true positive (TP), false positive (FP), true negative (TN), and false negative (FN) as follows: sensitivity (TP/(TP+FN) and specificity (TN/(TN+FP). (16)

### Measurements

Case ascertainment (stage 2) was completed on January 31, 2025, which represents the final date of the prevalence study. We defined the crude prevalence rate of PD as the number of persons identified as having PD divided by the total population studied (n = 31,009), then reported per 100,000 of the population. The data were sub-stratified by sex. Direct age-standardization was performed using both the WHO 2000 – 2025 World Standard Population (17) and the 2013 revision of the European Standard Population. (18) The age-standardized data were calculated using the automated Public Health England Analytical Tools for Public Health calculator. (17, 19) Age categories were in 5-year intervals, with the first and last collapsed (18 – 24 years and ≥85 years) for the convenience of computation, while retaining the sub-total population denominators inclusive of all ages within any category.

### Statistical methods

All survey data were captured electronically using Microsoft Excel ® spreadsheets created from the electronic questionnaires specifically designed for the study. Data were analyzed using IBM® Statistical Package for Social Sciences (SPSS®) Statistics version 26. Crude prevalence rates were calculated by dividing the number of PD cases by the total population screened overall and by sex and reported per 100,000 population. We performed direct age standardization from the Public Health England algorithm containing the embedded standardization formulae detailed in the spreadsheets associated with the Association of Public Health Observatories (APHO) Technical Briefing on Commonly Used Public Health Statistics and their Confidence Intervals. (20) In brief, rates are automatically calculated by imputing the local observed events (number of PD cases), local population size (number of screened participants in the age bracket), and the reference population (from the WHO World Standard Population tables and the 2013 revision of the European Standard Population) against each 5-year age stratum. (17–19) In this study, we computed the population size starting from age 18 and above and collapsed the first age stratum to 18 – 24, and the last age stratum to 85 and above. The directly standardized rates are reported with corresponding 95% confidence intervals. Similar calculations were conducted using sex-specific counts and population sizes available for the WHO World Standard Population. Demographic data for the study participants are reported using conventional summary statistics, and, where relevant, inter-group comparisons are provided using appropriate statistics (ANOVA for means and the chi-square test for proportions). The level of significance was set at a p-value <0.05. Sensitivity and specificity of the screening questionnaire was determined as described previously.

## RESULTS

### Main findings

The process of screening and inclusion/exclusion of participants from the final prevalence counts of persons with PD in this study is depicted in Figure 2. This includes the alternative primary diagnosis, excluding participants as not having PD. Stage 1 (household census and screening) included 13,222 unique households within which 31,009 eligible participants were individually screened. Basic demographics of the screened participants are shown in Table 1. The age distribution of participants and the corresponding number of prevalent PD cases overall and by sex strata are shown in Tables 2 and 3. 70 participants received a final diagnosis of PD, yielding a crude prevalence rate of 225.7 per 100,000 overall (17/16,351 (104.0) in women and 53/14658 (361.6) in men). The age-standardized prevalence rate (95% CI) based on the WHO World Standard Population was 193 per 100,000 (150 – 245) (women: 86 (50 – 137); men: 277 (207 – 362)). The age-standardized prevalence rate (95% CI) based on the 2013 Revision of the European Standard Population was 329 per 100,000 (249 – 424).

**Figure 2.**
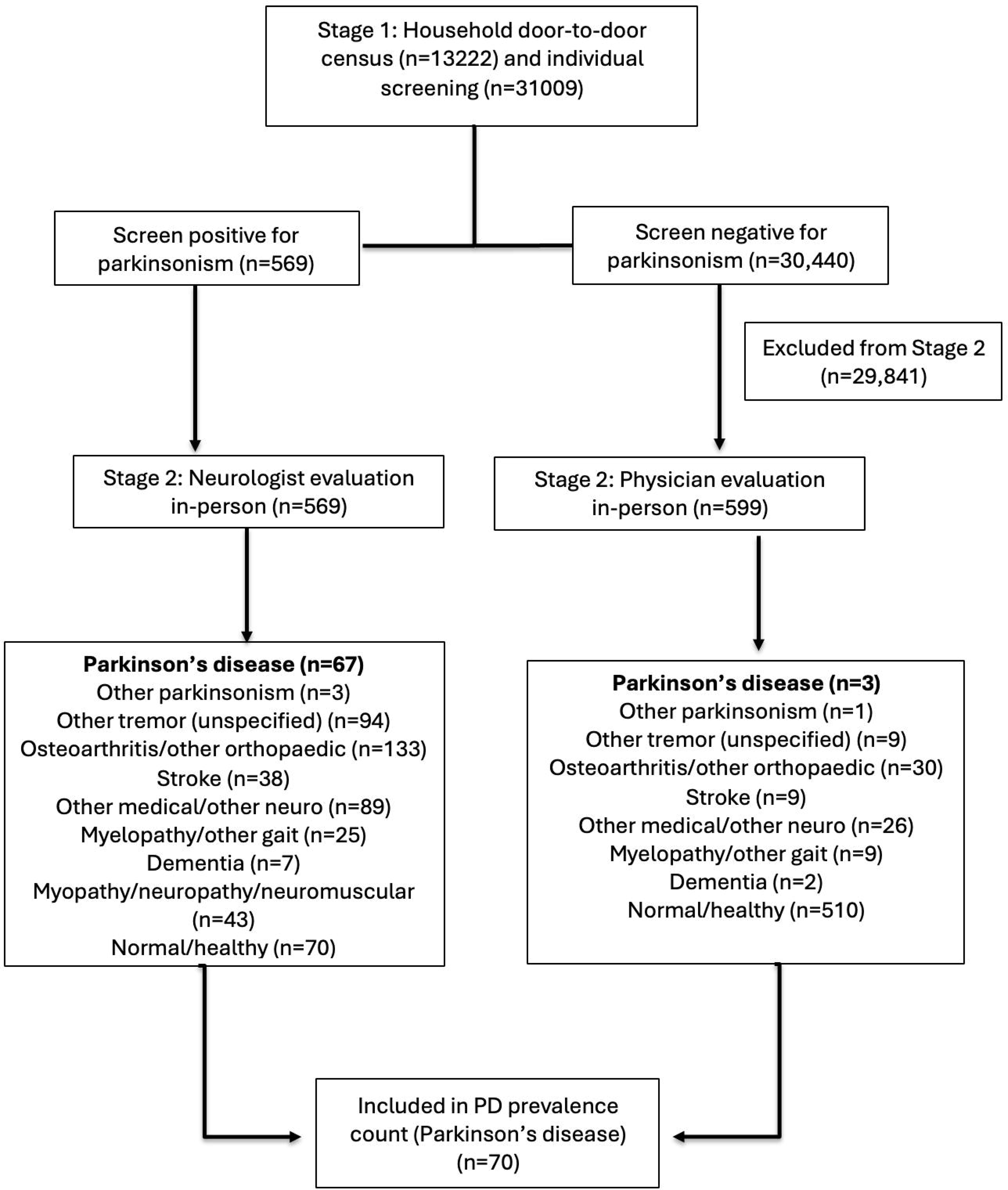
Participant identification flowchart

**Table 1.**
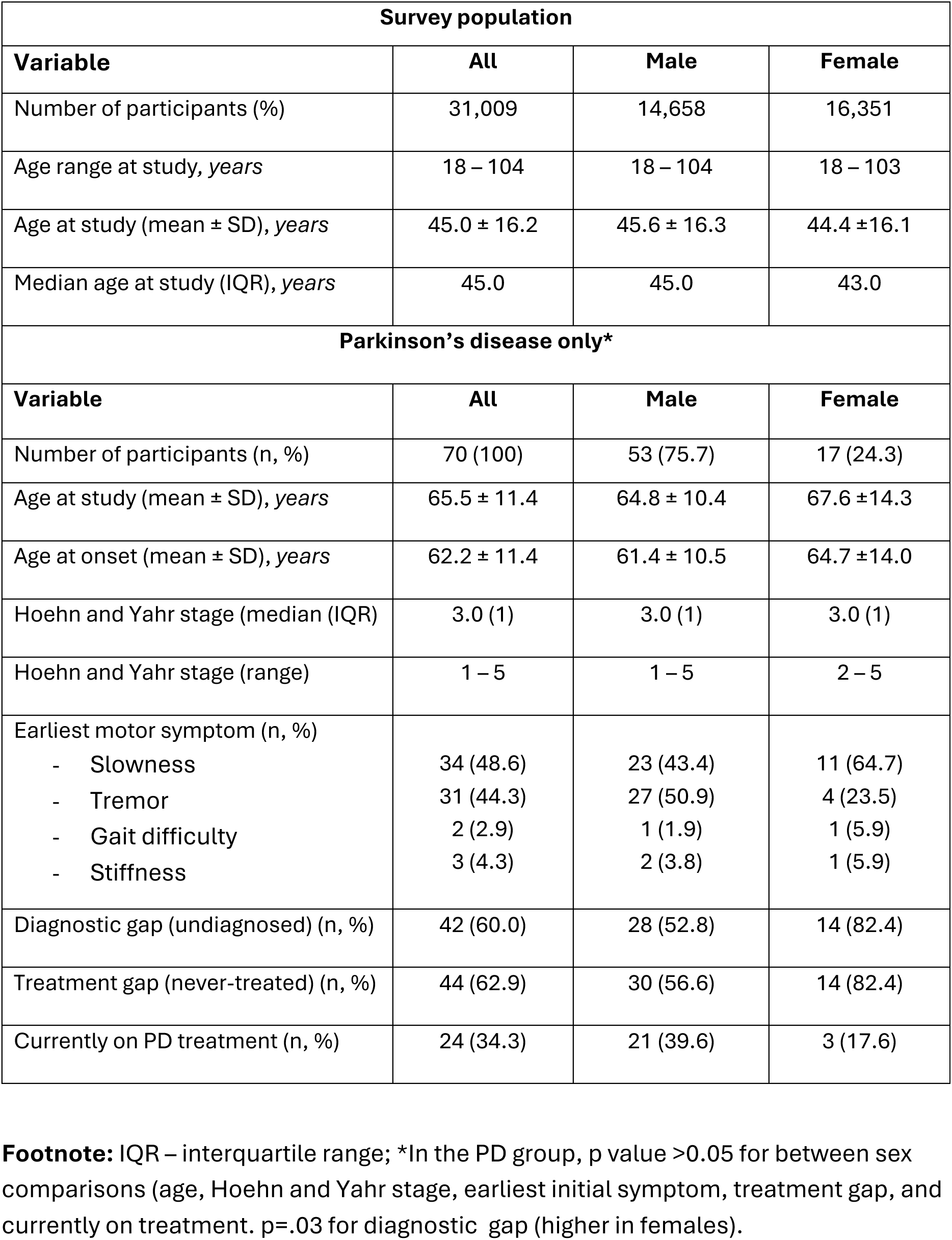
Demographic and clinical characteristics of the study population and persons with PD.

| Survey population |  |  |  |
| --- | --- | --- | --- |
| Variable | All | Male | Female |
| Number of participants (%) | 31,009 | 14,658 | 16,351 |
| Age range at study, *years* | 18 – 104 | 18 – 104 | 18 – 103 |
| Age at study (mean ± SD), *years* | 45.0 ± 16.2 | 45.6 ± 16.3 | 44.4 ± 16.1 |
| Median age at study (IQR), *years* | 45.0 | 45.0 | 43.0 |
| Parkinson's disease only* |  |  |  |
| Variable | All | Male | Female |
| Number of participants (n, %) | 70 (100) | 53 (75.7) | 17 (24.3) |
| Age at study (mean ± SD), *years* | 65.5 ± 11.4 | 64.8 ± 10.4 | 67.6 ± 14.3 |
| Age at onset (mean ± SD), *years* | 62.2 ± 11.4 | 61.4 ± 10.5 | 64.7 ± 14.0 |
| Hoehn and Yahr stage (median (IQR)) | 3.0 (1) | 3.0 (1) | 3.0 (1) |
| Hoehn and Yahr stage (range) | 1 – 5 | 1 – 5 | 2 – 5 |
| Earliest motor symptom (n, %) |  |  |  |
| - Slowness | 34 (48.6) | 23 (43.4) | 11 (64.7) |
| - Tremor | 31 (44.3) | 27 (50.9) | 4 (23.5) |
| - Gait difficulty | 2 (2.9) | 1 (1.9) | 1 (5.9) |
| - Stiffness | 3 (4.3) | 2 (3.8) | 1 (5.9) |
| Diagnostic gap (undiagnosed) (n, %) | 42 (60.0) | 28 (52.8) | 14 (82.4) |
| Treatment gap (never-treated) (n, %) | 44 (62.9) | 30 (56.6) | 14 (82.4) |
| Currently on PD treatment (n, %) | 24 (34.3) | 21 (39.6) | 3 (17.6) |

**Footnote:** IQR – interquartile range; \*In the PD group, p value $>0.05$ for between sex comparisons (age, Hoehn and Yahr stage, earliest initial symptom, treatment gap, and currently on treatment. $p=.03$ for diagnostic gap (higher in females).

**Table 2.** Population age distribution, proportion with Parkinson’s disease, and age-standardized prevalence rates.

| **Age groups** | **Parkinson's disease** | **Study population** | **Reference population (WHO 2000 – 2025 World Standard Population)<sup>17</sup>** | **Reference population (2013 Revised European Standard Population)\*<sup>18</sup>** |
| --- | --- | --- | --- | --- |
| **18-24** | 0 | 3,654 | 11,580 | 8,200 |
| **25-29** | 0 | 3,068 | 7,927 | 6,000 |
| **30-34** | 0 | 2,292 | 7,607 | 6,500 |
| **35-39** | 0 | 2,851 | 7,147 | 7,000 |
| **40-44** | 2 | 3,531 | 6,588 | 7,000 |
| **45-49** | 1 | 3,703 | 6,038 | 7,000 |
| **50-54** | 8 | 3,043 | 5,368 | 7,000 |
| **55-59** | 14 | 2,568 | 4,548 | 6,500 |
| **60-64** | 9 | 2,068 | 3,719 | 6,000 |
| **65-69** | 10 | 1,728 | 2,959 | 5,500 |
| **70-74** | 11 | 1,122 | 2,209 | 5,000 |
| **75-79** | 7 | 727 | 1,520 | 4,000 |
| **80-84** | 5 | 363 | 910 | 2,500 |
| **≥85** | 3 | 291 | 635 | 2,500 |
| **Total** | 70 | 31,009 | 68,755 | 80,700 |
| **Age-standardized prevalence rate per 100,000 (95% CI)** |  |  | **193 (150 – 245)** | **329 (249 – 424)** |

**Footnote:** \*European Standard Population age-stratified counts are based on EU-27 plus EFTA countries 2011-2030 projections (Annexe F) in reference document. (18) Ages 18-24 and ≥85 populations strata collapsed for convenience.

**Table 3.** Population age distribution and proportion with Parkinson’s disease (by sex stratification)

| Age groups | Female |  | Male |  | Reference population (WHO World Standard Population)* |
| --- | --- | --- | --- | --- | --- |
|  | Parkinson's disease | Study population | Parkinson's disease | Study population |  |
| 18-24 | 0 | 1,970 | 0 | 1,684 | 11,600 |
| 25-29 | 0 | 1,661 | 0 | 1,407 | 8,000 |
| 30-34 | 0 | 1,272 | 0 | 1,020 | 6,000 |
| 35-39 | 0 | 1,578 | 0 | 1,273 | 6,000 |
| 40-44 | 2 | 2,037 | 0 | 1,494 | 6,000 |
| 45-49 | 0 | 1,956 | 1 | 1,747 | 6,000 |
| 50-54 | 1 | 1,487 | 7 | 1,556 | 5,000 |
| 55-59 | 2 | 1,290 | 12 | 1,278 | 4,000 |
| 60-64 | 1 | 1,018 | 8 | 1,050 | 4,000 |
| 65-69 | 3 | 838 | 7 | 890 | 3,000 |
| 70-74 | 2 | 544 | 9 | 578 | 2,000 |
| 75-79 | 3 | 341 | 4 | 386 | 1,000 |
| 80-84 | 2 | 195 | 3 | 168 | 500 |
| ≥85 | 1 | 164 | 2 | 127 | 500 |
| Total | 17 | 16,351 | 53 | 14,658 | 63,600 |
| Crude prevalence rate per 100,000 | 103.97 |  | 361.58 |  |  |
| Age-standardized prevalence per 100,000 (95% CI) | 86 (50 – 137) |  | 277 (207 – 362) |  |  |

**Footnote:** \*Population sizes assigned to each sex category are same per the WHO World Standard Population tables.(17) Crude prevalence rate overall: 225.7 per 100,000

### Sensitivity and Specificity of the 5-item parkinsonism screening questionnaire

In total, 569 participants screened positive (endorsing at least 1 of the 5 questions as being present). 599 who screened negative were also re-evaluated, and based on the Stage 2 evaluation, the screening outcomes were: true positives (67), false positives (502), true negatives (596) and false negatives (3). As such, the sensitivity (TP/(TP+FN) and specificity (TN/(TN+FP) of the 5-item screening questionnaire for parkinsonism were 95.7 (95% confidence interval (CI) 88.0 – 99.1) and 54.3 (95% CI 51.3 – 57.3), respectively.

### Clinical and treatment characteristics of PD cases

Demographic and clinical characteristics for the entire study population and for persons with PD (age at study, age at onset, earliest motor feature of PD, diagnostic gap (never diagnosed), treatment gap (never treated), and proportion currently on treatment for PD) are shown in Table 1. Overall, there was a trend of greater diagnostic and treatment gaps in females, with only the difference in diagnostic gap reaching statistical significance (p=.03).

## DISCUSSION

Estimates of the burden of PD in Africa have inevitably relied on sparse and outdated data, mostly published decades ago. (1–3, 13, 21, 22) The rapidly increasing burden of PD, predominantly driven by population ageing, demographic transitions and population expansion in SSA, make contemporary epidemiological data mandatory. Using a population-based, multistage screening and diagnostic framework, the present study provides age- and sex-stratified estimates of PD prevalence derived from a clearly delineated urban population in western Africa. The screening instrument had excellent sensitivity (95.7%) and an acceptable specificity, making it appropriate for the scenario (avoiding missed cases) and compensating for the lower specificity through the 2-stage approach of in-person case ascertainment of PD status by trained clinicians. The systematic review of screening questionnaires for parkinsonism conducted by Dahodwala and colleagues described the sensitivity and specificity of the 9 screening instruments included in their review as ranging from 48-100% and 26-100% respectively. (23) Specifically, when used in a community setting, the most widely deployed questionnaire (the Tanner questionnaire) had a sensitivity of 61-92% and a specificity of 29 – 92%. (23) Variations are typically attributable to methodological and setting-specific differences such as the study personnel deployed, inclusion criteria (e.g. previously diagnosed PD only), individual items included, and the disease severity in the cohort. (23, 24) The disaggregation of the data into granular 5-year groups (excluding the first and final strata combined for convenience) aligns with current recommendations and enables adjustment for population age structure to facilitate valid comparison with other epidemiological studies. (25)

In this study, we have documented, for the first time, age-standardized prevalence rates in the SSA population that mirror the current prevalence of PD reported from other populations. The prevalence (age-standardized to the WHO World Population (193 per 100,000 (95% CI 150 – 245)) and the revised European Standard Population (329 per 100,000 (95% CI 249 – 424)) differs remarkably from estimates reported for Nigeria (40 to <50 per 100,000) in the systematic analysis of data from 1990 – 2016 as part of the GBD Study 2016. (2) It aligns better with the global pooled prevalence of 151 per 100,000 (95% CI 119 - 188) and more recent temporal trends in prevalence (for two time periods, 2000 – 2009 (118 per 100,000) and 2010 – 2023 (381 per 100,000) reported by Zhu *et al* in their meta-analysis of temporal trends in PD prevalence between 1980 and 2023. (12) Survey methodology impacts on prevalence data, and studies utilizing active community screening, similar to our approach, have generally reported higher prevalence than those based solely on medical records or administrative data, because of the advantage of identification of undiagnosed cases in community -based studies.

The observed male predominance in prevalence and the progressive increase in case frequency with advancing age are consistent with the established demographic profile of PD. However, we found a higher sex difference in prevalence than is generally reported. The sex difference declined each decade and was about 0.6 above age 80.

The sex differences may reflect demographic differences in the age structure of females residing in our urban population, in addition to the potential effect of the lower number of females with PD identified in our community (given the wider confidence interval).

Pringsheim *et al* reported a significant (three-fold) difference in sex only for individuals aged 50 – 59 years old (41 in females versus 134 in males) in their 2014 systematic review that included 47 geographically diverse studies with a similar methodology to ours (door-to-door or random population survey and 2 stage evaluation comprised of screening questionnaire and in-person case ascertainment). (24) Possible hormonal influences have been postulated to explain some of the sex differences. (24) It remains unclear if survival differences exist between the sexes in PD in Nigeria. Global estimates based on observational cohorts and clinic-based studies are mixed, with some studies indicating significantly higher male mortality, or marginal differences (higher in males), or no sex differences in PD mortality. (26–32) However, a higher burden of motor morbidity and complications in women with PD may impact outcomes, including survival, and may contribute to the sex difference in prevalent cases of PD observed in the present study. (33) Importantly, the methodological approach extended beyond prevalence estimation to incorporate diagnostic and treatment gaps, which reveal significant sex differences, being wider in females. Sex disparities in access to healthcare exist in Africa, with women often bearing a greater brunt of the consequential poorer outcomes from non-communicable diseases in general. (34, 35) The significantly higher diagnostic gap and the trend of a wider treatment gap in females also suggest that the observed prevalence may reflect not only underlying disease occurrence, but also patterns of detection and access to care, and the intersection with sociocultural and health systems determinants of disease burden.

There are limitations to our study which warrant consideration when interpreting the findings. There is a possibility that we missed mild, early or less typical presentations of PD in the screening stage. However, using a single affirmative response to define a screen positive and deploying the screening of a proportionately large number of screen negative individuals to estimate the sensitivity and specificity of our approach provided clarity as to the magnitude of this limitation which is at most modest. We do not envisage that this would significantly impact on the final prevalence estimates.

Differential participation by sex could also contribute to the observed diagnostic gaps. We did have a representative and comparable number of participants of each sex in each age stratum, and the age strata mirror the urban population distribution by age and sex for the study location. As prevalence reflects both incidence and survival, the cross-sectional nature of the analysis precludes inference regarding disease onset or progression, and the data should be interpreted in that context when comparing estimates with other populations and when using these data to inform service planning. Finally, as the understanding of PD and the burden attributable to non-motor features become more apparent, we acknowledge that using the current PD clinical diagnostic criteria that is reliant on the emergence of motor features means that our data exclude the important, contemporary and more broadly defined PD that recognizes prodromal and biologically defined disease. (36)

Ultimately, prevalence data underpins evidence-based decision-making and ensures that resource allocation is proportionate to the magnitude of the disease burden, especially where resources are limited. Consequently, our data have health systems, policy and research implications. The clarification that PD prevalence mirrors that documented globally, including in high- and middle-income countries makes it imperative to prioritize PD in public health policies and urgently implement the strategies recommended to reduce its burden. (6, 37)

The diagnostic and treatment gaps reflect broader weaknesses in health systems for the care of neurological disorders, including poor awareness, weak referral pathways, and limited medicines and specialist access. System-level interventions to close the gaps are crucial. These include adopting integrated care models that incorporate task sharing and shifting to first-contact health care professionals (e.g. community health extension workers, primary care physicians and nurses, and community pharmacists), broadening public awareness through the media and grass-roots mobilization, strengthening referral systems, improving health insurance structures and medicine access, and enacting equity-based policies that ensure vulnerable groups are served. (6, 38) Our data will contribute to future global disease burden estimates that will provide more credible and contemporary estimates for western Africa. Future research that employs longitudinal designs to distil the effects of incidence, survival, and care access on PD prevalence is recommended. In addition, repeat surveys or sentinel surveillance can track trends and evaluate policy impact.

## Supporting information

TraPCAf Consortium

## Data Availability

All data produced in the present work are contained in the manuscript

## ACKNOWLEDGMENTS

This study was funded by the National Institute for Health and Care Research (NIHR) in the UK (Award No. NIHR133391). The funding body provided peer review prior to awarding the research grant but did not play a role in the design of the study, nor will they have a role in the proposed data collection, management, analysis or interpretation of data.

---

**Supplementary Table 1.** Crude prevalence rates (%) per decade and sex stratification in study population.

| Age groups | Female |  |  | Male |  |  | All |  |  |
| --- | --- | --- | --- | --- | --- | --- | --- | --- | --- |
|  | PD | Study population | CP per 100 | PD | Study population | CP per 100 | PD | Study population | CP per 100 |
| **18-29** | 0 | 3631 | 0.00 | 0 | 3091 | 0.00 | 0 | 6722 | 0 |
| **30-39** | 0 | 2850 | 0.00 | 0 | 2293 | 0.00 | 0 | 5143 | 0 |
| **40-49** | 2 | 3993 | 0.05 | 1 | 3241 | 0.03 | 3 | 7234 | 0.04 |
| **50-59** | 3 | 2777 | 0.11 | 19 | 2834 | 0.67 | 22 | 5611 | 0.39 |
| **60-69** | 4 | 1856 | 0.22 | 15 | 1940 | 0.77 | 19 | 3796 | 0.50 |
| **70-79** | 5 | 885 | 0.56 | 13 | 964 | 1.35 | 18 | 1849 | 0.97 |
| **≥80** | 3 | 359 | 0.84 | 5 | 295 | 1.69 | 8 | 654 | 1.22 |
| **Total** | **17** | **16,351** | **0.10** | **53** | **14,658** | **0.36** | **70** | **31009** | **0.23** |

**Footnote:** Frequency of PD in population aged ≥60 years: 0.71% (45/6299) (0.39% in females (12/3100) and 1.03% in males (33/3199)).
CP - crude prevalence

